# Aerobic Exercise Training Rejuvenates the Human Skeletal Muscle Methylome Ten Years after Breast Cancer Treatment and Survival

**DOI:** 10.1101/2022.09.12.22279705

**Authors:** Piotr P. Gorski, Truls Raastad, Max Ullrich, Daniel C. Turner, Jostein Hallén, Sebastian Imre Savari, Tormod S. Nilsen, Adam P. Sharples

**Author notes:** Joint corresponding authors Tormod S. Nilsen -, Adam P. Sharples –.

## Abstract

Cancer survivors suffer impairments in skeletal muscle (SkM) in terms of reduced mass and function. Interestingly, human SkM possesses an epigenetic memory of earlier stimuli, such as exercise. Long-term retention of epigenetic changes in SkM following cancer survival and/or exercise training have not yet been studied. We therefore investigated genome-wide DNA methylation (methylome) in SkM following a 5-month, 3/week aerobic training intervention in breast cancer survivors 10-14 years after diagnosis and treatment. These results were compared to breast cancer survivors who remained untrained and to age-matched controls with no history of cancer, who undertook the same training intervention. SkM biopsies were obtained before(pre) and after(post) the 5-month training period and InfiniumEPIC 850K DNA methylation arrays performed. The breast cancer survivors displayed a significant retention of increased DNA methylation (i.e., hypermethylation) at a larger number of differentially methylated positions (DMPs) compared with healthy age-matched controls pre-training. Training in cancer survivors led to an exaggerated number of DMPs with a hypermethylated signature occurring at random non-regulatory regions across the DNA compared with training in healthy age-matched controls. However, the opposite occurred in important gene regulatory regions, where training in cancer survivors elicited a considerable reduction in methylation (i.e., hypomethylation) in 99% of the DMPs located in CpG islands within promoter regions. Importantly, training was able to reverse the hypermethylation identified in cancer survivors back towards a hypomethylated signature that was observed pre-training in healthy age-matched controls at 300 (out of 881) of these island/promoter associated CpGs. Pathway enrichment analysis identified training in cancer survivors evoked this predominantly hypomethylated signature in pathways associated with: Cell cycle, DNA replication/repair, transcription, translation, mTOR signalling and the proteosome. Differentially methylated region (DMR) analysis also identified genes: BAG1, BTG2, CHP1, KIFC1, MKL2, MTR, PEX11B, POLD2, S100A6, SNORD104 and SPG7 as hypermethylated in breast cancer survivors, with training reversing these CpG island/promoter associated DMRs towards a hypomethylated signature. Training also elicited a largely different epigenetic response in healthy individuals than that observed in cancer survivors, with very few overlapping changes. Only one gene, SIRT2, was identified as having altered methylation in cancer survivors at baseline as well as after training in both the cancer survivors and healthy controls. In conclusion, human SkM muscle retains a hypermethylated signature as long as 10-14 years after breast cancer treatment and survival. Five months of aerobic training rejuvenated the SkM methylome towards signatures identified in healthy age-matched individuals in gene regulatory regions.

## Introduction

In 2019, the World Health Organisation (WHO) suggested that cancer was ranked the first or second leading cause of death before 70 years of age in 112 (out of 183) countries [1]. Cancer incidents are also expected to increase to 28.4 million by 2040, which is a 47% increase compared to 2020 [1]. Breast cancer was the most diagnosed cancer in 2020 [1], surpassing lung cancer which was ranked as most highly diagnosed in 2018 [2]. This meant that in 2020 there were around 2.3 million new cases of breast cancer in women, accounting for 11.7% of all new cancer cases [1]. Breast cancer can be fatal if untreated or detected late with a mortality rate of approximately 15% [2]. However, modern treatments combining surgery, chemotherapy, hormone inhibition and/or radiotherapy means that most breast cancer patients will survive. For example, while breast cancer was the most common cancer worldwide in 2020 it was responsible for only 6.9% of cancer-related deaths compared with lung cancer, that accounted for 18% of cancer-related deaths [1]. However, the intense breast cancer therapy may also lead to late effects such as reduced aerobic capacity and increased risk of cardiovascular disease (CVD) [3-6]. It has been suggested that women, 5-6 years after breast cancer survival, have a 1.77-fold higher risk of CVD mortality compared to women without a history of breast-cancer [4]. Furthermore, breast cancer survivors have reduced SkM size, function, and performance (such as strength and mobility) [7, 8] that can contribute to increased morbidity and reduced quality of life. Aerobic exercise training can significantly increase aerobic capacity [9-11] and reduce the risk of CVD in breast cancer survivors [10], reviewed in [12]. Furthermore, aerobic exercise can improve SkM function, metabolism and performance which in turn are also associated with reduced CVD and morbidity as well as increased quality of life [11, 13-15].

Importantly, the effects of cancer diagnosis and treatment have been associated with accelerated ageing [16]. One of the mechanisms that may be responsible for this is epigenetic alterations that occur to DNA and chromatin, that are retained over time. This is because epigenetic modifications to DNA and the surrounding chromatin can be long-lasting after being exposed to different environmental stressors. In human SkM, we have previously demonstrated that DNA methylation is one of the key mechanisms responsible for ‘muscle memory’ as altered methylation profiles can be retained following both positive stimuli such as exercise training in humans [17, 18] and mice [19], as well as negative stimuli, such as high levels of inflammation [20] or high fat diets [21]. However, long-term retention of DNA methylation in SkM following cancer treatment and survival or epigenetic changes following exercise training in cancer survivors has not yet been studied.

DNA methylation has also been shown to form an accurate epigenetic clock that can predict both chronological and biological age [22-24]. Indeed, epigenetic age using DNA methylation from whole blood has been shown to increase in breast cancer patients undergoing treatment [25]. A recent study investigating childhood cancer survivors suggested that these individuals demonstrated accelerated epigenetic ageing into adulthood [26]. This is because, with age, it is proposed that our cells (including SkM [27-29]) accumulate methylation [22, 30] and therefore become ‘hypermethylated’. Meta-analysis of the SkM methylome also identified that hypermethylation predominantly occurs in gene regulatory regions with advancing age [31, 32]. In contrast, both acute exercise and chronic training have been shown to decrease methylation (i.e., hypomethylation) in both human and mouse SkM [17, 18, 33-35]. Therefore, exercise and increased physical activity are proposed to be epigenetically anti-ageing in SkM [28, 29, 36-39]. However, the role of exercise training on the ability to rejuvenate the epigenetic landscape and reduce the acceleration of epigenetic ageing in SkM after cancer survival is currently unknown.

Overall, we therefore aimed to 1) Investigate whether there is an altered genome-wide epigenetic landscape (DNA methylome) in SkM of breast cancer survivors, 10-14 years post diagnosis and treatment, with the hypothesis that there will be retained/accumulated profiles of DNA methylation compared with healthy (non-cancer) age-matched controls. 2) Investigate the influence of 5 months aerobic exercise training on the DNA methylome in SkM of breast cancer survivors, with the hypothesis that aerobic training will ‘rejuvenate’ the epigenetic landscape back towards DNA methylation signatures observed in healthy age-matched controls. 3) Investigate whether cancer survival affects an individual’s epigenetic age and whether exercise can alter epigenetic ageing using a recently developed muscle-specific epigenetic clock [31, 32]. For this, we hypothesise that cancer survival will increase epigenetic age and that aerobic training could prevent this increase when compared to cancer survivors who do not undertake training.

## Methods

### Ethical approval

This study is part of the ongoing CAUSE-trial (Cardiovascular Survivor Exercise) initiated by the Norwegian School of Sport Sciences (NSSS) and Oslo University Hospital. The Regional Committees for Medical and Health Research Ethics (REK) approved the study (reference number 28930; 2019/1318), and the study was pre-registered in clinicaltrials.gov (NCT04307407).

### Participants and Experimental Design

The CAUSE-trial is a two-armed randomised controlled exercise training trial, where breast cancer survivors are randomised to either an exercise group or a control group (usual care), that also includes third arm of age-matched women with no prior history of cancer undertaking the same exercise training program. Breast cancer survivors diagnosed, with stage II-III HER2 negative breast cancer at the age of 60 years or less, between 2008 and 2012 were identified by the Cancer Registry of Norway. The study included survivors that had received anthracycline-based chemotherapy (Epirubicin), were currently exercising less than 90 minutes at moderate to high intensity per week, and that were living within driving distance from NSSS. Major exclusion criteria for the study were stage IV breast cancer diagnosis, adjuvant Trastuzumab-treatment, recurrent breast cancer or presence of secondary cancers, previous major cardiac surgery, pacemaker, chronic atrial fibrillation or any recent or uncontrolled cardiovascular disease. Participants for the healthy age-matched control group were recruited via advertisements. The participants were matched for age with the breast cancer survivors and could not have a history of any cancer and otherwise had to adhere to the same eligibility criteria as the breast cancer survivors (except for the breast cancer diagnosis).

The breast cancer survivors were block randomised with a 1:1 allocation ratio to either an aerobic exercise training group (*Cancer Trained*) or to a control group (*Cancer Untrained*) for 5 months (20 weeks). The training intervention is described below. Participants in the healthy age-matched group did not undergo randomization, but all underwent the same exercise protocol as the cancer trained group (*Healthy Age-Matched Trained*). All participant groups had SkM biopsies taken and underwent other study-related assessments pre and post training. SkM biopsy methods are detailed below. For this study, pre and post muscle biopsies were analysed from 7, 6 and 10 participants in the Cancer Trained, Cancer Untrained and Healthy Age-Matched Trained group, respectively. Baseline characteristics of age, height, body mass and BMI for all participants are outlined in **Table 1**, with no difference between groups.

**Table 1.** Baseline characteristics of the participants

|  | **Cancer**<br>**Trained**<br>**(n=7)** | **Cancer**<br>**Untrained**<br>**(n=6)** | **Healthy Age-**<br>**Matched**<br>**Trained**<br>**(n=10)** | **Survivors vs.**<br>**Healthy Age-**<br>**Matched (p-**<br>**values)** |
| --- | --- | --- | --- | --- |
| Age (year; SD) | 62.5 (4.2) | 59.9 (5.7) | 57.6 (4.0) | 0.089 |
| Body Height (cm; SD) | 167.8 (2.79) | 171.3 (5.4) | 169.3 (5.0) | 0.854 |
| Body Mass (Kg; SD) | 81.7 (4.7) | 77.6 (14.4) | 77.2 (15.0) | 0.670 |
| Body Mass Index (SD) | 29.1 (2.2) | 26.5 (4.9) | 26.9 (5.1) | 0.691 |
| $\dot{V}O_{2peak}$ (ml/kg/min; SD) | 26.2 (4.2) | 29.0 (7.4) | 30.2 (6.3) | 0.339 |
| Physical activity (PA) levels<br>(minutes; SD) |  |  |  |  |
| Light PA | 152.2 (39.3) | 152.1 (26.4) | 158.1 (29.7) | 0.162 |
| Moderate to Vigorous PA | 41.5 (25.4) | 45.6 (34.5) | 62.4 (27.9) | 0.163 |

### Aerobic Exercise Training

Both the cancer survivors training group and the healthy age-matched group underwent five months of 3 times per week treadmill-based endurance training, aimed at increasing 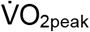. The program followed a non-linear periodization model, typically with one hard (i.e., 80-87% of HR_peak_), one moderate (i.e., 70-80% of HR_peak_) and one easy (i.e., 60-70% of HR_peak_) session per week. Initially, all sessions were continuous in duration, but from week 5, the hard session was an interval session, with 3-5 intervals each lasting 4 minutes (i.e., 90-97% of HR_peak_). After week 10, we introduced 8-minute intervals for the moderate session (i.e., 85-91% of HR_peak_), progressing from 2-4 intervals throughout the intervention.

### Cardiopulmonary Exercise Test and Physical Activity levels

Participants body height and mass were recorded using a stadiometer and a scale (SECA 213, Hamburg, Germany). Cardiorespiratory fitness 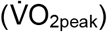 was assessed by a treadmill-based, symptom-limited cardiopulmonary exercise test, using a stepwise modified Balke Protocol until exhaustion [40]. After familiarisation with the treadmill, participants were encouraged to walk at gradually increasing exercise loads (increased inclination or speed) until voluntary exhaustion. Continuous expired air was measured breath-by-breath, using a gas and volume calibrated metabolic chart (Oxycon Pro, Jarger GmbH, Hoechberg, Germany). Objectively measured physical activity levels were recorded for seven consecutive days, using the ActiGraph(tm) model GT3X+ (ActiGraph LLC, Pensacola FL, USA). The average minutes spent in light and moderate-to-vigorous physical activity, using cut-off values used previously [41], with light physical activity defined as 100-1951 counts per minute and moderate-to-vigorous physical activity >1952 counts per minute. Baseline characteristics of 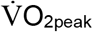 and physical activity levels for all participants can be found in **Table 1**. There was no significant difference in 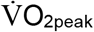 or physical activity levels between the cancer groups and the healthy age-matched group prior to the start of the exercise program.

### Skeletal Muscle Biopsies

The biopsy procedure was conducted under local anaesthesia (Xylocaine with adrenaline, 10 mg/ml lidocaine + 5 µg/ml adrenaline, AstraZeneca, London, UK) and muscle tissue was obtained using a modified Bergström technique with suction. The tissue was quickly rinsed in physiological saline, before fat, connective tissue and blood were removed and discarded. Subsequently, the samples were quickly frozen in isopentane and cooled on dry ice or liquid nitrogen, before being transferred and stored at −80°C for later isolation of DNA and RNA. All biopsies from both pre and post training time points were taken at rest, in the morning after an overnight fast from the lateral portion of vastus lateralis muscle. All participants were asked only to consume water during the overnight fast and therefore did not consume alcohol and caffeine the night before or the morning of the pre and post aerobic training biopsy. The participants were also asked not to perform any exercise two days before the pre training biopsy. Post training samples were obtained at least 72 hrs after the end of the training intervention. Pre and post biopsies were taken from the left leg in all participants.

### Tissue Homogenization, DNA isolation, Bisulfite Conversion and Genome-Wide DNA Methylation

DNA was isolated from SkM tissue derived from a randomly selected subpopulation within each group (n=5 cancer trained, n=5 cancer untrained and n=6 healthy age-matched trained) at both pre and post training. These subpopulations baseline characteristics can be found in **Table 2**. Baseline characteristics were well balanced between the groups (**Table 2**). As was the case in the entire cohort, described above (**Table 1**). There was no difference in age, BMI, 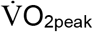 or physical activity levels between the cancer groups and the healthy age-matched group. Muscle samples were homogenized for 45 seconds at 6,000 rpm × 3 (5 min on ice in-between intervals) in lysis buffer (180 µl buffer ATL with 20 µl proteinase K) provided in the DNeasy spin column kit (Qiagen, UK) using a Roche Magnalyser instrument and homogenization tubes containing ceramic beads (Roche, UK). The DNA was then bisulfite converted using the EZ DNA Methylation Kit (Zymo Research, CA, United States) as per the manufacturer’s instructions.

**Table 2.** Baseline characteristics for the subpopulation of DNA isolated for genome-wide DNA methylation analysis.

|  | **Cancer**<br>**Trained**<br>**(n=5)** | **Cancer**<br>**Untrained**<br>**(n=5)** | **Healthy**<br>**Age-**<br>**Matched**<br>**Trained**<br>**(n=6)** | **Survivors**<br>**vs. Healthy**<br>**Age-**<br>**matched (p-**<br>**values)** |
| --- | --- | --- | --- | --- |
| Age (year; SD) | 60.4 (5.9) | 62.0 (5.5) | 58.3 (4.8) | 0.307 |
| Body Height (cm; SD) | 167.5 (5.9) | 168.1 (5.0) | 169.0 (4.8) | 0.660 |
| Body Mass (Kg; SD) | 80.5 (7.9) | 74.1 (8.6) | 80.0 (16.5) | 0.670 |
| Body Mass Index (SD) | 28.8 (3.8) | 26.4 (4.3) | 28.0 (5.8) | 0.861 |
| $\dot{V}O_{2peak}$ (ml/kg/min; SD) | 27.2 (3.9) | 30.9 (6.7) | 30.7 (7.7) | 0.630 |
| Physical activity (PA) levels (minutes; SD) |  |  |  |  |
| Light PA | 154 (44.9) | 154 (38.4) | 162.9 (28.3) | 0.652 |
| Moderate to Vigorous PA | 43.3 (34.6) | 41.8 (16.4) | 56.2 (31.6) | 0.362 |

### Infinium MethylationEPIC BeadChip Array

All DNA methylation experiments were performed in accordance with Illumina manufacturer instructions for the Infinium Methylation EPIC BeadChip Array. Methods for the amplification, fragmentation, precipitation and resuspension of amplified DNA, hybridisation to EPIC beadchip, extension and staining of the bisulfite converted DNA (BCD) can be found in detail in our open access methods paper [17, 33]. EPIC BeadChips were imaged using the Illumina iScan System (Illumina, United States).

### DNA methylome analysis, differentially methylated Positions (DMPs), pathway enrichment analysis (KEGG pathways) and differentially methylated region (DMR) analysis

Following MethylationEPIC BeadChip arrays, raw .IDAT files were processed using Partek Genomics Suite V.7 (Partek Inc. Missouri, USA) and annotated using the MethylationEPIC_v-1-0_B4 manifest file. The mean detection p-value for all samples was 0.0004 (**Suppl. Figure 1a**), which was well below the recommended 0.01 in the Oshlack workflow [42]. We also examined the raw intensities/signals of the probes, that demonstrated an average median methylated and unmethylated signal of 11.77 and 11.69, respectively, with an average of 11.73, which is recommended to be above 11.5 [42]. The difference between the average median methylated and average median unmethylated signal was 0.07, well below the recommended difference of less than 0.5 [42]. Upon import of the data into Partek Genomics Suite we filtered out probes located in known single nucleotide polymorphisms (SNPs) and any known cross-reactive probes using previously defined SNP and cross-reactive probe lists from EPIC BeadChip 850K validation studies [43]. Although the average detection p-value for each sample across all probes was very low (on average 0.0004) we also excluded any individual probes with a detection p-value that was above 0.01 as recommended previously [42]. Furthermore, due to an all-female cohort of breast cancer survivors and age-matched healthy controls, we filtered out any probes on the male only Y chromosome (therefore the analysis included all probes on the X chromosome only). Out of a total of 865,860 probes in the EPIC array, removal of known SNPs, cross-reactive probes, those with a detection p-value above 0.01 and those on the Y chromosome resulted in 808,804 probes being taken forward for downstream analysis. Following this, background normalisation was performed via functional normalisation (with noob background correction) as previously described [44]. After functional normalisation, we also undertook quality control procedures via principal component analysis (PCA) (**Suppl. Figure 1b**). One sample (sample 13, **Suppl. Figure 1b**) was removed due to a larger variation than that expected within that condition (variation defined as values above 2.2 standard deviations (SDs) for that condition – depicted by ellipsoids in the PCA plots -**Suppl. Figure 1b**). Following normalisation and quality control procedures, we undertook differentially methylated position (DMP) analysis by converting β-values to M-values (M-value = log2(β / (1 - β)), as M-values show distributions that are more statistically valid for the differential analysis of methylation levels [45]. We then performed a two-way ANOVA for group/condition (cancer trained, cancer untrained, healthy age-matched trained) and time (pre and post), with planned contrast/pairwise comparisons of: Cancer Trained-Pre vs. Healthy Trained-Pre (to investigate the effect of Cancer on the methylome named - *Cancer Alone*), Cancer Trained-Post vs. Cancer Trained-Pre (to investigate the impact of training on the methylome of breast cancer survivors named - *Training in Cancer*), Healthy Age-Matched Trained-Post vs. Heathy Age-Matched-Pre (to investigate the impact of training on the methylome in age-matched healthy controls named-*Training in Healthy*), Cancer Untrained-Post vs. Cancer Untrained-Pre (to help assess if any methylation changes occur over the time of the intervention period in cancer survivors, named - *Experimental Time*), and finally Cancer Trained Pre vs. Cancer Untrained Pre (to assess if there were any differences in methylation between the cancer groups at baseline, named - *Between Cancer Groups at Baseline*). For initial discovery of CpG sites that were deemed statistically significant, DMPs with an unadjusted P value of ≤ 0.01 were accepted for downstream analysis (Kyoto Encyclopedia of Genes and Genomes/KEGG pathway, differentially methylated region/DMR analysis - see below). We then undertook CpG enrichment analysis on these DMPs within KEGG pathways [46-48] using Partek Genomics Suite and Partek Pathway software. DMR analysis was performed to identify where several CpGs were differentially methylated within a short chromosomal locations/regions, undertaken using the Bioconductor package DMRcate (DOI: <u>10.18129/B9.bioc.DMRcate</u>).

### Removal of confounding differential DNA methylation over experimental time and starting variation between cancer survivor groups at baseline

To investigate the influence of 5 months aerobic training on DNA methylation in cancer survivors compared with healthy age-matched controls, we first wished to remove any differential methylation that was observed without training in cancer survivors over time (the 5-month intervention period). Therefore, using a relevant independent control group of cancer survivors who performed no training (Cancer Untrained group) during the 5-month period, we could ascertain and remove any differential methylation that was influenced by experimental time. There were 1,325 differentially methylated positions (DMPs) that were altered over the experimental time of 5 months. However, only 108 out of these 1,325 DMPs were influenced by aerobic training in either the cancer survivors or healthy age-matched controls (**Figure 1A**). Indeed, this was only 0.6% of the DMPs (90 out of 14,645 DMPs) that were affected by training in cancer survivors and 1.05% of DMPs (23 out of 2,195 DMPs) that were affected by training in healthy age-matched controls (**Figure 1A**). We therefore removed these 108 DMPs from any downstream analysis so that any differential methylation observed to be altered with training was due to the training intervention and not a consequence of the experimental time-period. Furthermore, while all cancer survivors in the study had previously been diagnosed and treated for breast cancer 10-14 years earlier, prior treatment programmes were often individualised and different. Therefore, to account for any potential variation in starting (baseline) methylation profiles, we sought to identify if there was any differential methylation between the two cancer survivor groups at baseline (i.e., Cancer Trained Pre vs. Cancer Untrained Pre) to remove any differences in DNA methylation that could have been influenced by starting variation between cancer survivors before the intervention period. Indeed, there were 6,860 DMPs identified between cancer survivor groups at baseline. However, only 378 out of these 6,860 DMPs were influenced by aerobic training in both the cancer survivors and healthy age-matched controls (**Figure 1B**). Indeed, this was only 2.4% of the DMPs (356 out of 14,645 DMPs) that were affected by training in cancer and 1.14% of DMPs (25 out of 2,195 DMPs) that were affected by training in healthy age-matched controls (**Figure 1B**). Therefore, in addition to the confounding experimental time DMPs (108 DMPs) removed above, we also removed the 378 DMPs identified at baseline between the cancer survivor groups, so that any downstream discovery of differential methylation would be due to the training intervention rather than having been influenced by starting differences that may occur in methylation between cancer survivors. This meant removal of 469 DMPs in total, as 17 of the same confounding DMPs identified with experimental time (108 DMPs) were also identified between cancer groups (378 DMPs).

**Figure 1.**
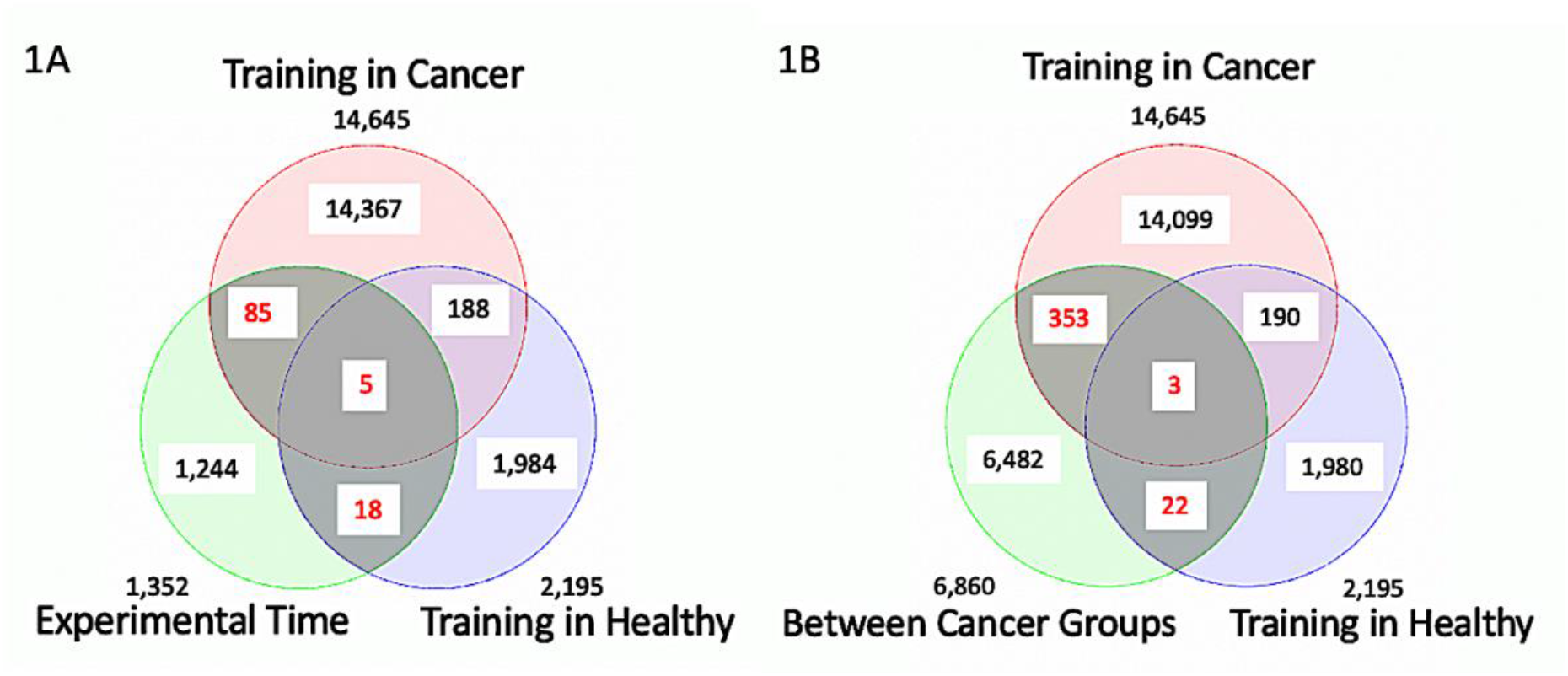
**(A)** There were 1,325 differentially methylated positions (DMPs) that were altered over the experimental time of 5 months. Only 108 out of these 1,325 DMPs were influenced by aerobic training in either the cancer survivors or healthy age-matched controls (overlapping numbers in red font). These 108 DMPs were removed from later analysis so that any differential methylation observed to be altered with training was due to the training intervention and not a consequence of the experimental time period of the training intervention. **(B)** 6,860 DMPs identified between cancer survivor groups at baseline. Only 378 out of these 6,860 DMPs were influenced by aerobic training in both the cancer survivors and healthy age-matched controls (overlapping numbers in red font). Therefore, we also removed these 378 DMPs so that any downstream discovery of differential methylation would be due to the training intervention rather than having been influenced by starting differences in methylation between cancer survivor groups.

### Muscle-specific epigenetic clock and age estimation

Ageing is associated with DNA methylation changes in all human tissues, and epigenetic markers can estimate chronological age based on DNA methylation patterns [22]. Together with others we have recently helped developed a muscle-specific epigenetic clock (muscle-specific epigenetic age test, or ‘MEAT’) that predicts age with better accuracy than the pan-tissue epigenetic clock [31, 32]. Therefore, for the analysis of DNA methylation age (DNAm Age), we extracted the β-values from normalized data, as previously described. Then we reduced the size of the dataset to only contain the 18,747 probes / CpGs that were common between the 17 datasets and the 1,053 samples used to create the MEAT clock [31, 32] that estimates DNAm Age using the MEAT2.0 elastic net regression model. The DNAm Age estimations were calculated with the R package version MEAT2.0 using R 4.1.3. Finally, we also calculated age acceleration or deviation (AA) which is DNAm Age – chronological age, also the difference in age acceleration after the training (post) compared with before training (pre) abbreviated as AAdiff post-pre. A positive age acceleration occurs when DNAm-Age is higher than chronological age and individuals are said to have ‘accelerated epigenetic aging’ and a negative age acceleration occurs when chronological age is higher than DNAm-Age and individuals are said to have ‘decelerated epigenetic aging’.

### RNA isolation, primer design & gene expression analysis

Muscle tissue was homogenised in tubes containing ceramic beads (MagNA Lyser Green Beads, Roche, Germany) and 1 ml Tri-Reagent (Invitrogen, UK) for 45 seconds at 6,000 rpm × 3 (and placed on ice for 5 min at the end of each 45 second homogenization) using a Roche Magnalyser instrument (Roche, Germany). RNA was then isolated as per Invitrogen’s manufacturer’s instructions for Tri-reagent. A one-step RT-qPCR reaction (reverse transcription and PCR) was performed using QuantiFast SYBR Green RT-PCR one-step kits on a Rotorgene 3000Q thermocycler (Qiagen, UK). Each reaction was setup as follows; 4.75 μl experimental sample (4.21 ng/μl totalling 20 ng per reaction), 0.075 μl of both forward and reverse primer of the gene of interest (100 μM stock suspension), 0.1 μl of QuantiFast RT Mix (Qiagen, Manchester, UK) and 5 μl of QuantiFast SYBR Green RT-PCR Master Mix (Qiagen, Manchester, UK). Each sample was run in duplicate. Reverse transcription was initiated with a hold at 50°C for 10 min (cDNA synthesis) and a 5 min hold at 95°C (transcriptase inactivation and initial denaturation), before 45 × PCR cycles of; 95°C for 10 sec (denaturation) followed by 60°C for 30 sec (annealing and extension). Primer sequences for genes of interest and reference genes are in **Table 3**.

**Table 3.**
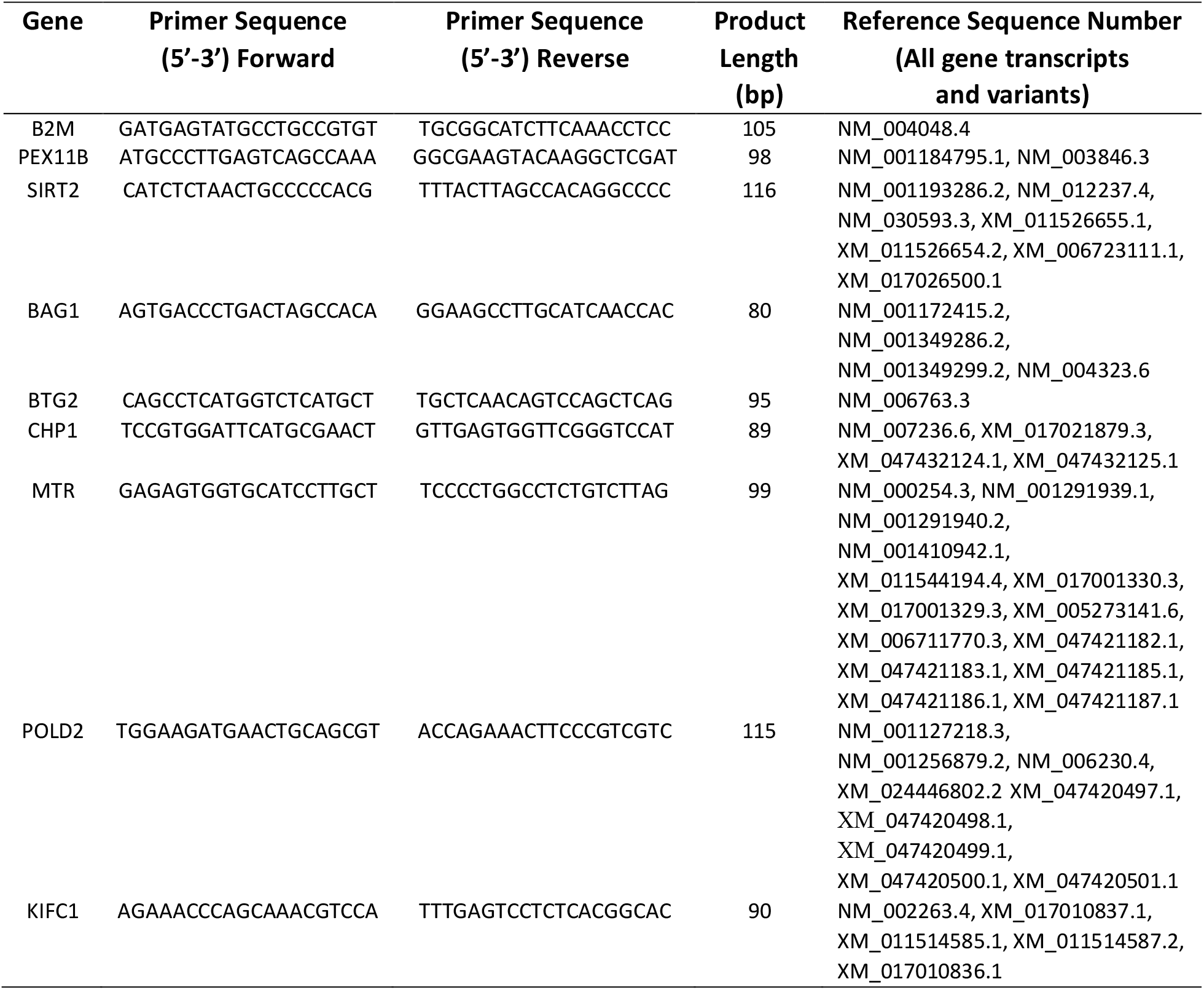
Primers details for RT-qPCR.

All primers were designed to yield products that included the majority of transcript variants for each gene as an impression of total changes in the gene of interests’ expression levels. All genes demonstrated no unintended gene targets via BLAST search and yielded a single peak after melt curve analysis conducted after the PCR step above. All relative gene expression was quantified using the comparative Ct (^ΔΔ^Ct) method [49]. The baseline sample for each participant and their own reference gene sample was used as the calibrator in the cancer trained pre and post comparison. For the baseline (pre) comparison between healthy age-matched controls and cancer trained group at baseline (pre) the pooled Ct value of pre healthy age-matched controls were used as calibrator. The average, standard deviation and variations in Ct value for the B2M reference gene demonstrated low variation across all samples (mean ± SD, 16.82 ± 0.54, 3.23% variation) for the analysis. The average PCR efficiencies for the genes of interest were comparable (average of 89.92 ± 2.45%) with the reference gene B2M (89.90 ± 1.58%). However, KIFC1 and BAG1 had higher amplification values 107.35 ± 16.99% and 102.93 ± 28.70% respectively, while the melt curves yielded a single peak. Due to lack of RNA available we were unable to perform further analysis on KIFC1 and BAG1 and we include the data for these two genes with this caveat in mind. Gene expression and statistical analysis was performed on n = 5 to 10 individuals per condition in duplicate (see figure legends for precise details). For the cancer alone comparison (Healthy Age-Matched Trained-Pre vs. Cancer Trained-Pre), a two-tailed unpaired t-test at the level of p ≤ 0.05 was performed. For the gene expression analysis for training in cancer condition (Cancer Trained-Pre vs. Cancer Trained-Post) a two-tailed paired t-test at the level of P value of ≤ 0.05 was performed. For analysis of SIRT2 gene expression pre and post training in both the healthy age-matched trained and cancer trained group was analysed using a two-way ANOVA with Tukey post-hoc tests. Statistical analysis was performed on GraphPad Prism (version 9.2.0).

## Results

### Hypermethylation of the skeletal muscle epigenome occurs after treatment and survival of breast cancer 10 -14 years previously

The SkM tissue of cancer survivors demonstrated 6,102 significant DMPs compared to healthy age-matched controls at baseline (pre-training), 10-14 years after breast cancer treatment and survival. Therefore, signifying a large number of CpG sites differentially methylated between cancer survivors and healthy age-matched controls (**Suppl. File 1A**). Indeed, the DNA methylation in SkM of cancer survivors demonstrated a more hypermethylated signature, with 57% of DMPs being hypermethylated and 43% being hypomethylated (3,500 vs. 2,602 DMPs, respectively) compared with healthy age-matched controls (**Figure 2A**). Importantly, 881 DMPs were in CpG islands within gene promoter regions, and the proportion of hypermethylation in CpG islands within promoter regions of cancer survivors was even more profound, with 98% (866 DMPs) hypermethylated compared with only 2% (15 DMPs) hypomethylated **Figure 2B** & **2C; Suppl. File 1B**. The hypermethylation in CpG islands within promoters was enriched most notably in relevant pathways of: Ubiquitin Mediated Proteolysis, Cell Cycle, Mitophagy, Autophagy and p53 pathways (see **Figure 2D** and **2E, Suppl. File 1C**). Furthermore, there were many genes demonstrating differential methylation in short chromosomal regions (or differentially methylated regions known as DMRs) in cancer survivors. There were 262 DMRs identified between cancer survivors and healthy age-matched controls at baseline (pre-training) (**Suppl. File 1D**) and 46 of these DMRs were in CpG islands within promoters (**Suppl. File 1E**). The majority of these DMRs in islands within promoters (43 out of 46) demonstrated hypermethylation (i.e., a positive mean beta difference of the DMPs contained within that DMR) in cancer survivors compared with healthy age-matched controls (**Suppl. File 1E**).

**Figure 2.**
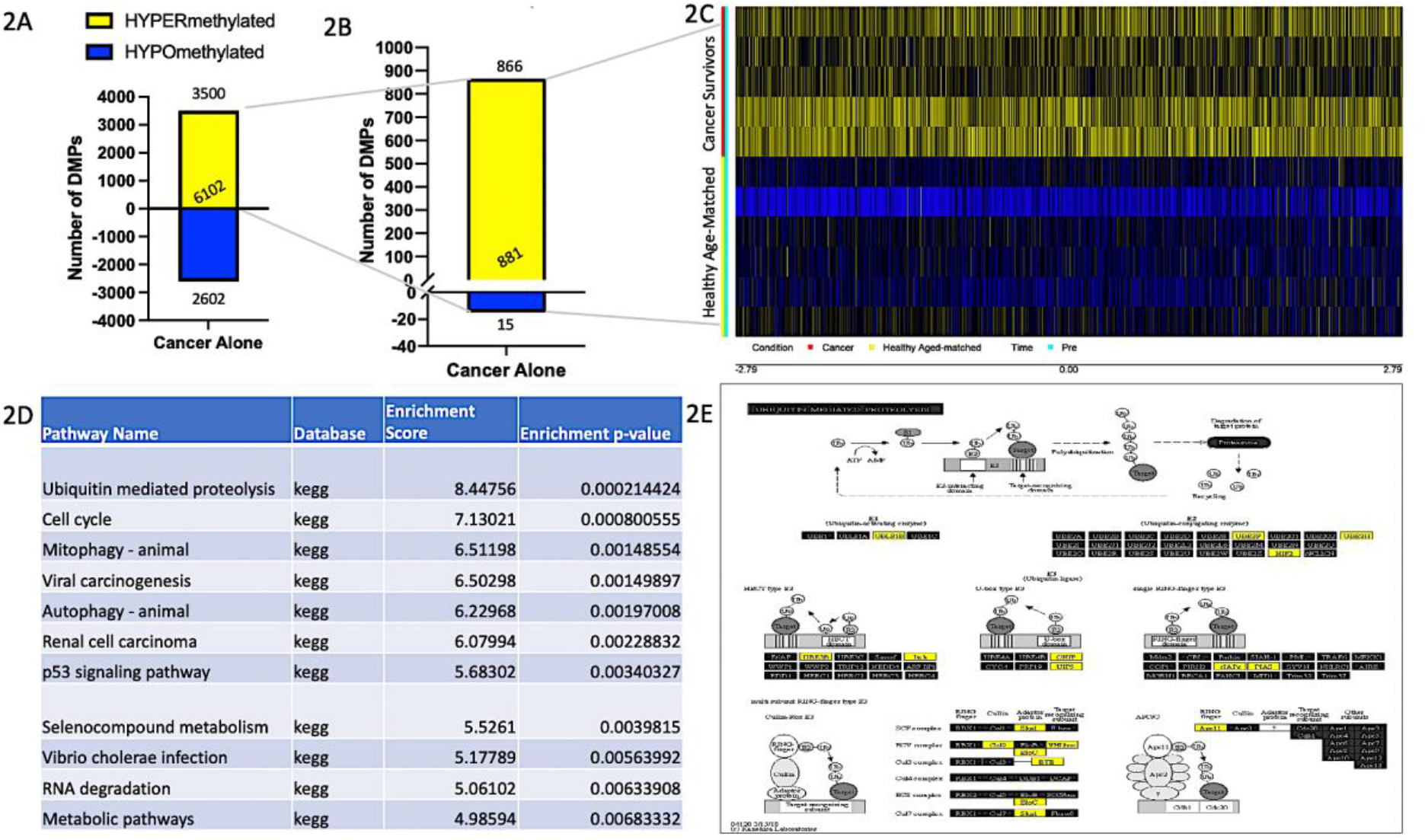
**(A)** Demonstrates the number of all differentially methylated positions (DMPs) in the SkM of breast cancer survivors compared with healthy age-matched controls (cancer alone) (57% hypermethylated in yellow and 43% of DMPs hypomethylated in blue). **(B)** When interrogating gene regulatory regions, 98% of DMPs located in CpG islands within promoters (866 out of 881 DMPs) were hypermethylated in cancer survivors compared with healthy age-matched controls. **(C)** Heatmap depicting the 881 DMPs located in CpG islands within gene promoters and their methylation signature showing predominantly a hypermethylated (yellow) profile in cancer survivors compared with hypomethylated (blue) in healthy age-matched controls at baseline/ pre-training. **(D)** Table of the top enriched KEGG pathways between cancer survivors and healthy age-matched controls at baseline [46-48]. **(E)** Pathway image of top ranked KEGG pathway – Ubiquitin mediated proteolysis in cancer survivors compared with healthy age-matched controls. Yellow coloured genes depict hypermethylated genes within this pathway. Note that this KEGG pathway image is generated using the most significant CpG site / DMP within each gene and therefore when there is more than one CpG site per gene these may not be representative of all changes within an individual gene. Therefore, the image is simply a representative image of the most significant methylation changes within the genes of this pathway. However, as suggested in the results, 98% of the DMPs used to generate the pathway image were hypermethylated in cancer survivors compared with healthy age-matched controls. Consequently, it is likely that even where genes possessed more than one CpG site the methylation status would have been the same and the genes would have been coloured the same in the pathway image. For the accurate list of all 881 DMPs used for the KEGG pathway analysis see **Suppl. File 1B** and specifically the DMPs used to generate the ubiquitin mediated proteolysis pathway image see **Suppl. File 1C** column Z onwards.

### Aerobic exercise training in cancer survivors evokes a profound impact on the DNA methylome of skeletal muscle and reverses the methylome signature seen after cancer survival towards a profile observed in healthy age-matched controls

Training in cancer survivors evoked a more profound impact on DNA methylation across the genome compared with healthy age-matched controls **(Figure 3A)**. Where, 14,215 CpG sites were differentially methylated after training in cancer survivors **(Suppl. File 2A)** compared with 2,149 DMPs after training in healthy age-matched controls **(Suppl. File 2B)**. Across the genome, training evoked predominantly a hypermethylated signature in both cancer survivors (8,586 hyper vs. 5,629 hypo-methylated DMPs) and healthy age-matched controls (1,745 hyper vs. 404 hypomethylated DMPs, see **Figure 3A and 3B**). However, the hypermethylation that occurred with training was predominantly located in regions without a regulatory feature. Yet, when interrogating differential methylation of gene regulatory regions of CpG islands within promoter regions, this switched to a more hypomethylated signature. Therefore, training in both cancer and healthy participants preferentially evoked hypomethylation within CpG islands located in promoter regions. With training in cancer survivors evoking a larger absolute hypomethylating stimulus (2,358 hypo-vs. 2 hyper-methylated) corresponding to 99.9% of DMPs being hypomethylated in islands within promoter regions in cancer survivors (**Figure 3C and 3D, Suppl. File 2C)**, compared to healthy age-matched controls generating a smaller absolute hypomethylating stimulus (124 sites / 82% hypomethylated vs. 23 sites / 18% hypermethylated; **Suppl. File 2D**).

**Figure 3.**
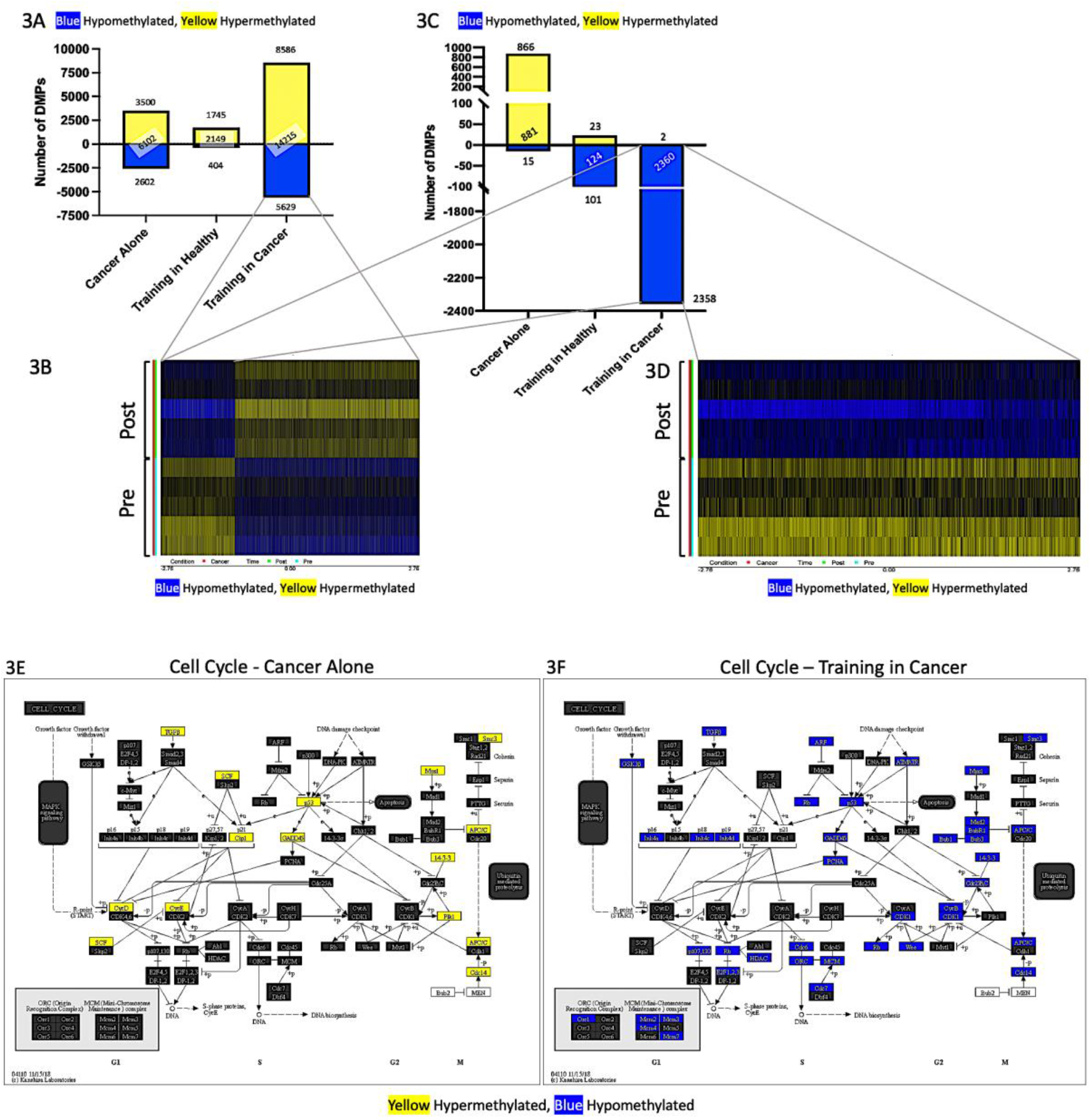
**(A)** Demonstrates the number of all differentially methylated positions (DMPs) in the SkM of breast cancer survivors compared with healthy age-matched controls (cancer alone) as well as all differentially methylated positions (DMPs) altered with training in healthy age-matched controls (Training in Healthy) and after training in cancer survivors (Training in Cancer). **(B)** Heatmap depicting all 14,215 DMPs and their methylation signature (hypomethylated blue, hypermethylated yellow) before (pre) and after (post) training in cancer survivors. **(C)** DMPs located in islands and promoters after cancer survival, and training in both healthy age-matched controls and cancer survivors. When interrogating DMPs within gene regulatory regions of CpG islands within promoters 99.9% (2,358 out of 2,360 DMPs) were hypomethylated after training in cancer survivors. **(D)** Heatmap depicting 2,360 DMPs in CpG islands within gene promoter regions demonstrating a hypomethylated signature post training, compared with pre training in cancer survivors. **(E)** Cell Cycle KEGG pathway image [46-48] demonstrating hypermethylation (yellow) in cancer survivors at baseline (pre-training) compared with healthy age-matched controls within this enriched pathway. **(F)** Cell Cycle KEGG pathway image post vs. pre training in cancer survivors, demonstrating the pathway becomes hypomethylated (blue) after training compared to hypermethylated (yellow) with cancer (E). Note that these KEGG images in 3E & F are generated using the most significant CpG site / DMP within each gene and therefore when there is more than one CpG site per gene these may not be representative of all changes within each individual gene. Therefore, the figure is simply a representative image of the most significant changes of the genes in this pathway. However, as suggested in the results 98% of the DMPs used in the pathway analysis for cancer survivors vs. healthy age-matched controls (E) were hypermethylated, and 99.9% of the DMPs used in the pathway analysis for post vs. pre training in cancer survivors were hypomethylated. Therefore, it is likely that even where genes possessed more than one CpG site the methylation status would have been the same and the genes would have been coloured the same in the pathway image. For the complete list of all 881 DMPs used for the KEGG pathway analysis in cancer alone conditions see **Suppl. File 1B** and specifically the DMPs in Cell Cycle pathway in cancer alone see **Suppl. File 1C (**column DB onwards). For the complete list of all 2,360 DMPs used for the KEGG pathway analysis in training in cancer conditions see **Suppl. File 2C** and specifically the DMPs in Cell Cycle pathway after training in cancer see **Suppl. File 2F**.

In cancer survivors, the hypomethylation in CpG islands within promoters was enriched in pathways associated with the Cell Cycle (top ranked KEGG pathway) and predominantly those involved in DNA replication, DNA excision repair, RNA transport and degradation, mTOR signalling and the proteosome (**Suppl. File 2E**). Indeed, as described above, the Cell Cycle pathway was also the second most highly ranked enriched pathway in cancer survivors compared to healthy age-matched controls at baseline, prior to any training (**Figure 2D** and **2E, Suppl. File 1C**). Where genes in the Cell Cycle pathway demonstrated predominantly a hypermethylated signature (**Figure 3E**), whereas, after training in cancer survivor’s genes in this pathway demonstrated a reversal of this trend with enrichment of hypomethylation within islands and promoters (**Figure 3E & F**). Overall, suggesting aerobic training can reverse the hypermethylation amongst cell cycle genes within SkM that occurs after cancer survival to a hypomethylated signature.

Given this reversal of methylation within islands and promoters within this top enriched pathway (Cell Cycle) after training in cancer survivors, we also confirmed that all the overlapping DMPs in islands and promoters that were affected by both cancer survival alone, and training in cancer (a total of 300 overlapping/ shared DMPs -**Figure 4A**) demonstrated a shift from hypermethylation with cancer survival to hypomethylation after training. Importantly, this represented a shift to a similar hypomethylated profile seen in healthy age-matched controls at baseline (**Figure 4B**). Overall, demonstrating that training 10-14 years after cancer survival rejuvenated the methylome in islands within promoters towards signatures observed in healthy age-matched individuals.

**Figure 4.**
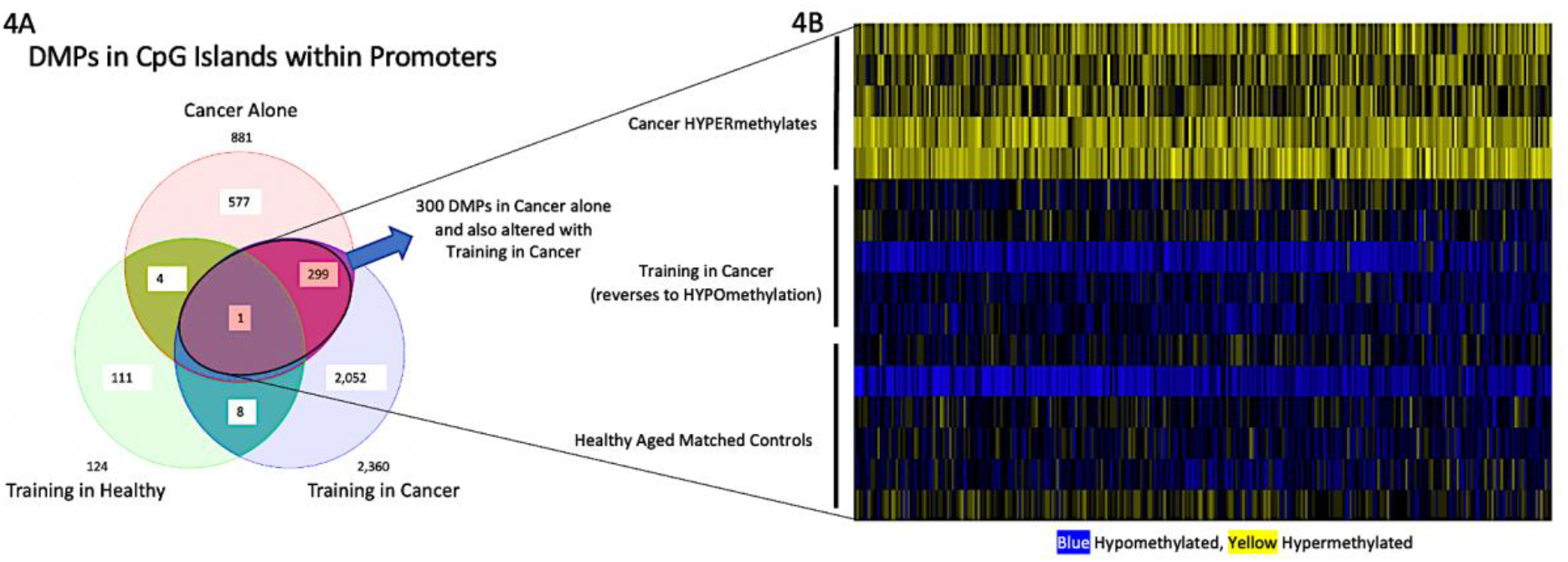
**(A)** Number of differentially methylated positions (DMPs) across conditions located in CpG islands within promoter regions. Three hundred DMPs were shared/overlapped (highlighted ellipse) between cancer survival (cancer alone) and after training in cancer survivors. **(B)** Heatmap depicts these 300 shared DMPs in islands within promoters that were altered with cancer survival alone and with training in cancer. After cancer survival alone, these DMPs were predominantly hypermethylated (yellow, top panel), then after training in these cancer survivors this profile was reversed to a hypomethylated state (blue, middle panel), which was similar to the hypomethylated profile seen in healthy age-matched controls at baseline /pre-training (blue, lower panel).

Furthermore, due to the rejuvenation of the methylome after training in cancer survivors, to identify which genes were most likely to be affected at their gene expression level we analysed differential methylation of short chromosomal regions, also known as differentially methylated regions or DMRs. Firstly, there were a large total number of genes with DMRs in cancer survivors after training, where we identified 972 DMRs (953 with annotated gene names -**Figure 5A, Suppl. File 2G**) with 298 of these DMRs located in CpG islands within promoter regions (**Suppl. File 2H**), where again all these DMRs were hypomethylated demonstrated by a negative mean beta difference for all DMPs within each DMR (**Suppl. File 2H**). As described earlier in the results, analysis of cancer survivors at baseline demonstrated 262 DMRs (260 in annotated genes) compared with healthy age-matched controls at baseline (pre training) (**Suppl. File. 1D**) with 46 of these DMRs located in islands within promoters (**Suppl. File 1E**). There were 43 DMRs that were altered in cancer alone and training in cancer (**Figure 5A**). Where 13 of these DMRs were in islands within promoter regions of genes: BAG1, BTG2, CHP1, KIFC1, MKL2, MTR, PEX11B, POLD2, RP11-467P9.1, RP11-649E7.5, S100A6, SNORD104, SPG7 (**Figure 5B**). Indeed, all these DMRs were hypermethylated with cancer survival compared with healthy age-matched controls at baseline (pre-training) (**Suppl. File 2I)** and were reversed to a hypomethylated profile after training in cancer survivors (**Suppl. File 2J)**. For example, the DMR with most CpG sites altered with training in cancer survivors, PEX11B, possessed a hypermethylated DMR with cancer survival alone (Chromosome 1:145516272 - 145516448) which was reversed to a hypomethylated state after training in cancer survivors in the same region (chromosome 1: 145516064 – 145516624), see **Figure 5C**.

**Figure 5.**
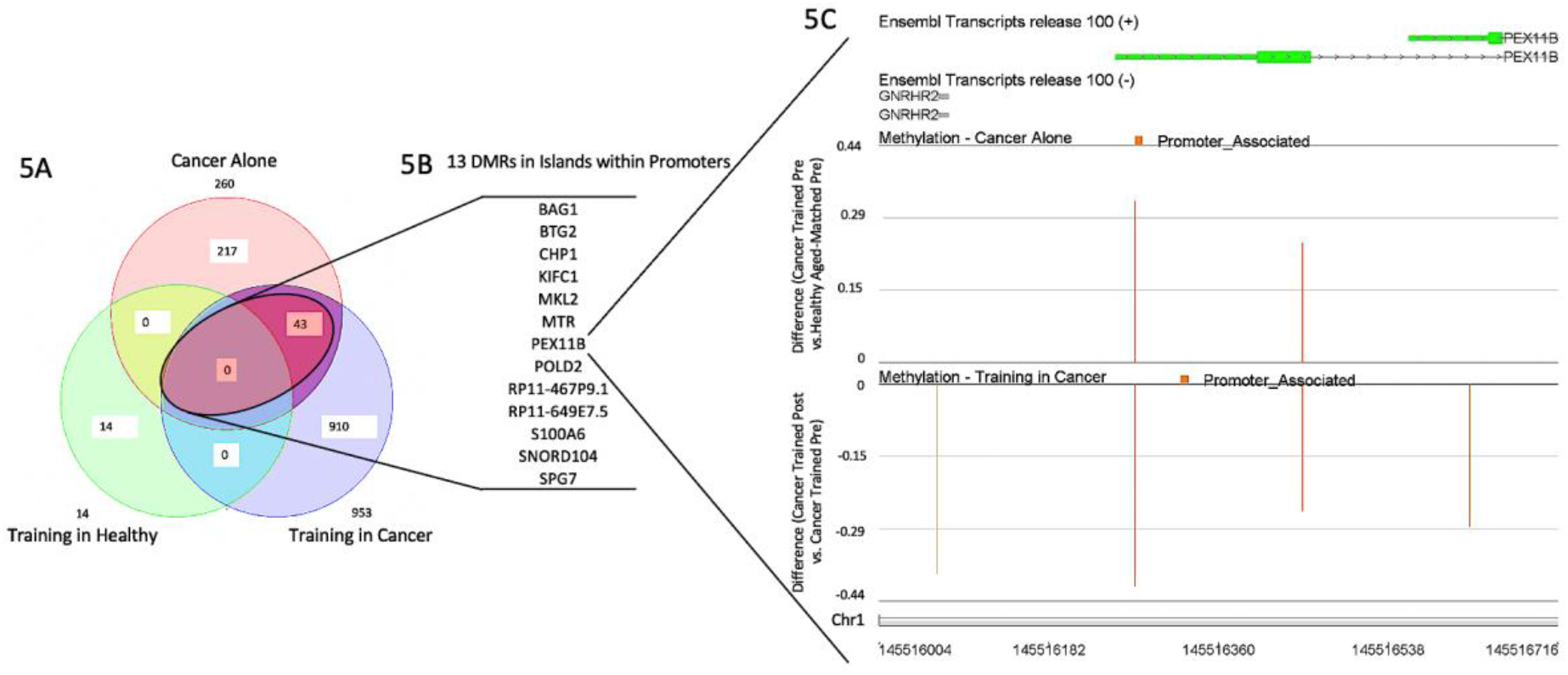
**(A)** Venn diagram of the overlap between the number of differentially methylated regions (DMRs) in the SkM of breast cancer survivors compared with healthy age-matched controls (cancer alone) and compared with the number of DMRs after training in healthy age-matched controls (training in healthy) and after training in cancer survivors (training in cancer). There were 260 DMRs annotated by gene name detected in SkM after cancer survival and 953 DMRs annotated by gene name identified after training in cancer survivors, with only 14 DMRs influenced by training in the healthy age-matched individuals. No DMRs that were altered after training in cancer were also altered after training in healthy individuals. There were 43 shared/overlapping DMRs that were affected with cancer alone that were also affected after training in cancer survivors. **(B)** 13 (out of 43) DMRs shared between cancer survival and after training in cancer survivors were in islands within promoter regions. All these DMRs were hypermethylated after cancer survival which were reversed to being hypomethylated after training in cancer survivors. **(C)** An example of the DMR for the promoter region of gene PEX11B. This gene was hypermethylated at 2 CpG sites after cancer survival that then became hypomethylated at the same positions after the training period, as well as 2 additional hypomethylated CpG sites in the same chromosomal region of the PEX11B promoter region.

Finally, because altered methylation at multiple CpG sites in a small region (DMR) located in islands within a promoter are more likely to affect gene expression, we ran the gene expression for some of the genes that possessed these DMRs (**Figure 6**). We ran 7 (out of 13) of these DMR genes (PEX11B, BTG2, CHP1, MTR, POLD2, KIFC1, BAG1), because RP11-467P9.1, RP11-649E7.5, S100A6 and SNORD104 were only on predicted transcripts, MKL2 gene had over 30 transcript variants making it difficult to design primers to detect changes in global gene expression and SPG7 primers demonstrated amplification of more than one product. We were able to demonstrate that together with these genes possessing hypermethylation in their promoter regions in cancer survivors when compared to healthy age-matched controls at baseline (described above), that 6 (out of 7) of these genes demonstrated an average reduction in gene expression in the cancer survivors, reaching statistical significance in 5 (out of 6) of these genes (PEX11B, MTR, POLD2, BAG1, KIFC1) (**Figure 6A-6E**), with an average but non-significant reduction in CHP1 (**Figure 6F**), with BTG2, significantly increasing in expression in the cancer trained group at baseline compared with the healthy age-matched controls (**Figure 6G**). As described above, aerobic training was able to hypomethylate these gene promoter regions, and on average, 4 (out of 7) of these genes demonstrated an increase in gene expression post training in the cancer group (MTR, POLD2, BAG1, KIFC1; **Figure 6H-K**), however, these were not significantly different. BTG2 however demonstrated a significant reduction in gene expression after training in the cancer survivors (**Figure 6L**).

**Figure 6.**
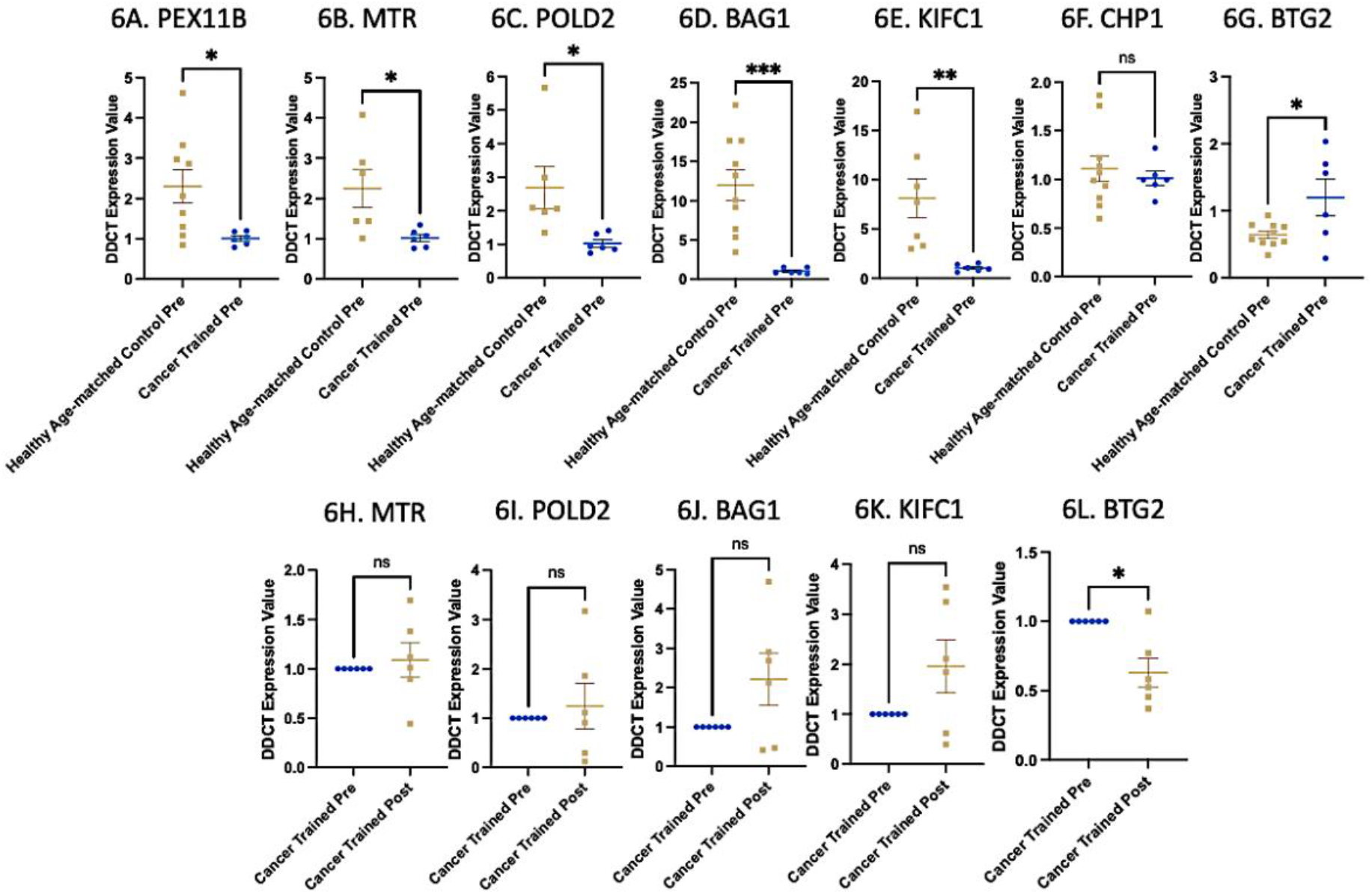
**A-G**. Gene expression of genes identified to have hypermethylated DMRs in cancer survivors compared with healthy age-matched controls. The majority of genes showing significant reductions in gene expression in cancer survivors, except BTG2 that significantly increased in expression in cancer survivors. Healthy age-matched controls n = 10 BAG1, CHP1 and BTG2, n = 9 PEX11B, n = 7 KIFC1, n = 6 MTR and POLD2. n = 6 all genes cancer trained pre. **H-L**. Gene expression of the genes that possessed hypomethylated DMRs after training in cancer and demonstrated average increases in gene expression. *p<0.05, ** p<0.005, *** p<0.0005, ns = not statistically significant. n = 6 cancer trained pre vs. cancer trained post.

### Training after cancer survival remodels the methylome in distinct genes and pathways compared to healthy individuals after the same 5-month training program

As described above both DMP and DMR analysis identified that cancer survivors demonstrated a hypermethylated DNA signature in islands within promoter regions and that training was able to reverse these signatures towards a hypomethylated signature of that observed in healthy age-matched controls. Also, as described above, training in healthy individuals did not evoke as considerable impact on the methylome as it did in cancer survivors. Where, 14,215 CpG sites were differentially methylated after training in cancer survivors **(Figure 7A, Suppl. File 2A)** compared with 2,149 DMPs after training in healthy age-matched controls **(Figure 7A, Suppl. File 2B)**. Therefore, it is worth highlighting that training in healthy age-matched controls differentially methylated predominantly different CpG sites in different genes and pathways than those modulated by e aerobic training in cancer survivors. Indeed, training in healthy individuals only affected 186 DMPs that were also altered by training in cancer survivors (**Figure 7A**). With only 9 DMPs located in islands within promoter regions that were altered after training in both cancer survivors and healthy individuals (**Figure 7B**). Despite this, these 9 DMPs (on 8 annotated genes SIRT2, NPHP4, IMMP2L, RBL2, PGRMC2, RAD51L3, TAF5, C3orf71) in islands within promoters may provide the only insight into aerobic training responsive DMPs that are independent of prior health/disease status.

**Figure 7.**
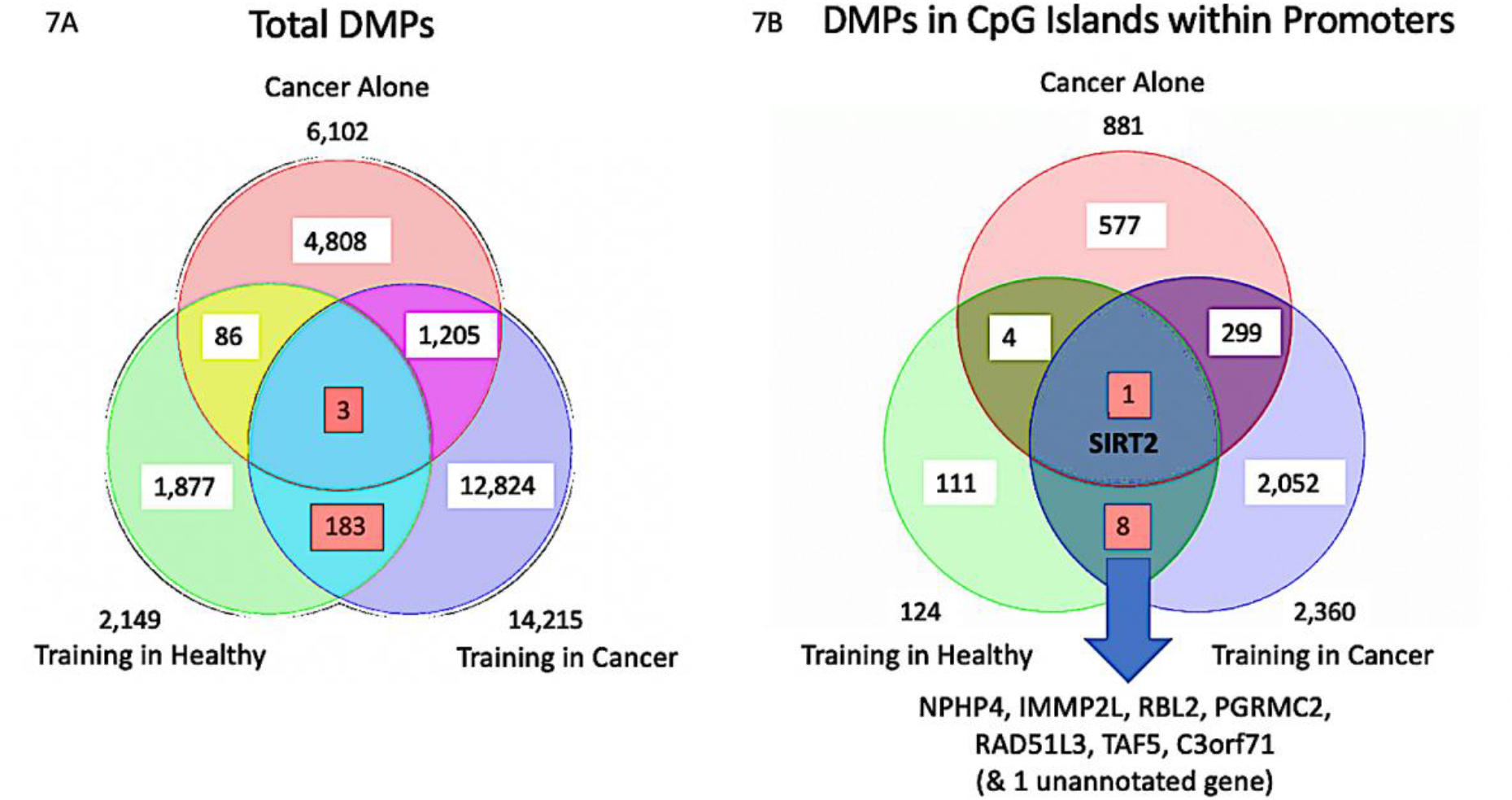
**(A)** Venn diagram of overlap between the number of all differentially methylated positions (DMPs) in the SkM of breast cancer survivors compared with healthy age-matched controls (Cancer Alone) and compared with the number of DMPs after training in healthy age-matched controls (Training in Healthy) and after training in cancer survivors (Training in Cancer). Training in healthy individuals did not evoke as considerable impact on the methylome as it did in cancer survivors. Where, 14,215 CpG sites were differentially methylated after training in cancer survivors compared with 2,149 DMPs after training in healthy age-matched controls. Training in healthy individuals only affected 186 DMPs that were also altered by training in cancer survivors (number highlighted in red boxes). **(B)** Venn diagram of overlap between the (DMPs) located in islands within promoter regions, where only 9 DMPs (on 8 annotated genes SIRT2, NPHP4, IMMP2L, RBL2, PGRMC2, RAD51L3, TAF5, C3orf71) located in islands within promoter regions were altered after both training in cancer survivors and healthy individuals (numbers highlighted in red boxes). The DMPs on these genes were therefore dependant on aerobic training and not dependant on prior disease/health status.

In further support of the divergent epigenomic response to training between cancer survivors and healthy individuals, there were only 14 DMRs discovered after training in healthy individuals (**Suppl. File 3A**) and only 1 located within an island within a promoter (**Suppl. File 3B**), compared to the large number of DMRs (972 with 953 on annotated genes) after training in cancer (with 298 of these DMRs located in islands within promoter regions-described above and already depicted in **Figure 5A**). Where importantly, none of these DMRs altered after training in cancer were altered after training in healthy individuals (**Figure 5A**). Therefore, there were no differentially methylated regions (DMRs) on CpG islands within promoter regions of genes that were affected by training in healthy individuals that were also affected by training in cancer survivors. Overall, suggesting that the epigenetic response to training in cancer survivors was substantially different than age-matched healthy controls.

### SIRT2 is an epigenetically responsive cancer and training gene

SIRT2 was the only gene identified to possess a DMP within a CpG island within its promoter region that was differentially methylated in cancer survivors at baseline and was also impacted by training in both cancer survivors and healthy age-matched controls (see **Figure 7B**). Indeed, SIRT2 (probe cg22546859) was hypermethylated in cancer survivors and then hypomethylated at the same site (probe cg22546859) as well as at another site (cg04614655) after training in cancer. However, it was oppositely hypermethylated (probe cg22546859) after training in healthy individuals. Interestingly, where SIRT2 was hypermethylated in cancer survivors at baseline compared with healthy age-matched controls, there was a reduction in its gene expression at the level of p = 0.0686 (**Figure 8A**). On average where there was hypomethylation of SIRT2, there was an average increase in gene expression after aerobic training in cancer survivors (**Figure 8B**), however these differences were not statistically significant. Furthermore, the healthy age-matched controls also had an average increase in SIRT2 gene expression that was also non-significant. Overall, SIRT2 gene is differentially methylated after both disease (cancer) and training regardless of disease history (i.e., in both cancer and healthy individuals) but in an alternate fashion in cancer survivors compared with healthy individuals. Cancer survivors also possessed reduced gene expression at baseline compared to healthy age-matched controls, however both groups demonstrated an average increase in SIRT2 gene expression that was not statistically significant.

**Figure 8.**
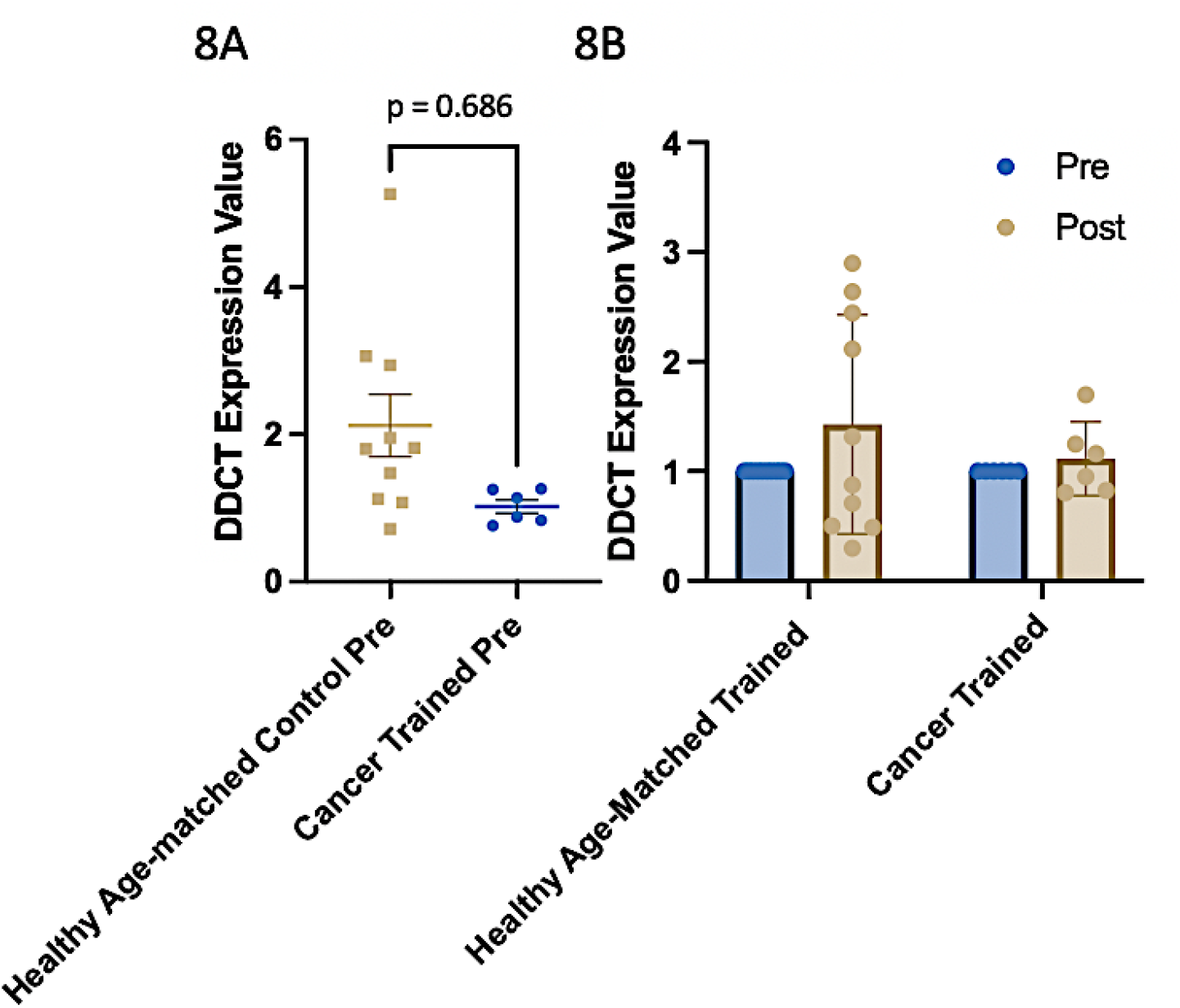
**A**. SIRT2 gene expression in cancer survivors compared with healthy age-matched trained controls at baseline/pre training. **B**. Gene expression of SIRT2 after training in both the healthy age-matched trained group and in the cancer trained group pre and post aerobic training. n = 10 healthy age-matched controls, n = 6 cancer trained.

### Epigenetic clock analysis reveals that cancer survival increases epigenetic muscle tissue age compared with healthy age-matched controls

Both cancer survival groups at baseline (cancer trained and cancer untrained) possessed higher DNAm Age compared with healthy age-matched controls (**Figure 9A**), suggesting a trend for increased epigenetic age in the muscle following cancer survival. However, this increase was not statistically significant. Interestingly, on average the epigenetic age of cancer survivors who did not train increased over the experimental time course. Whereas training in cancer survivors was able to prevent this average increase. Despite these trends, this was not statistically significant. Given this data, we also calculated DNA methylation age acceleration, where training in cancer survivors where there was a trend of reduce age acceleration (**Figure 9B**) that was not altered in the healthy age-matched controls or untrained cancer survivors. However, this difference was also not statistically significant between groups.

**Figure 9.**
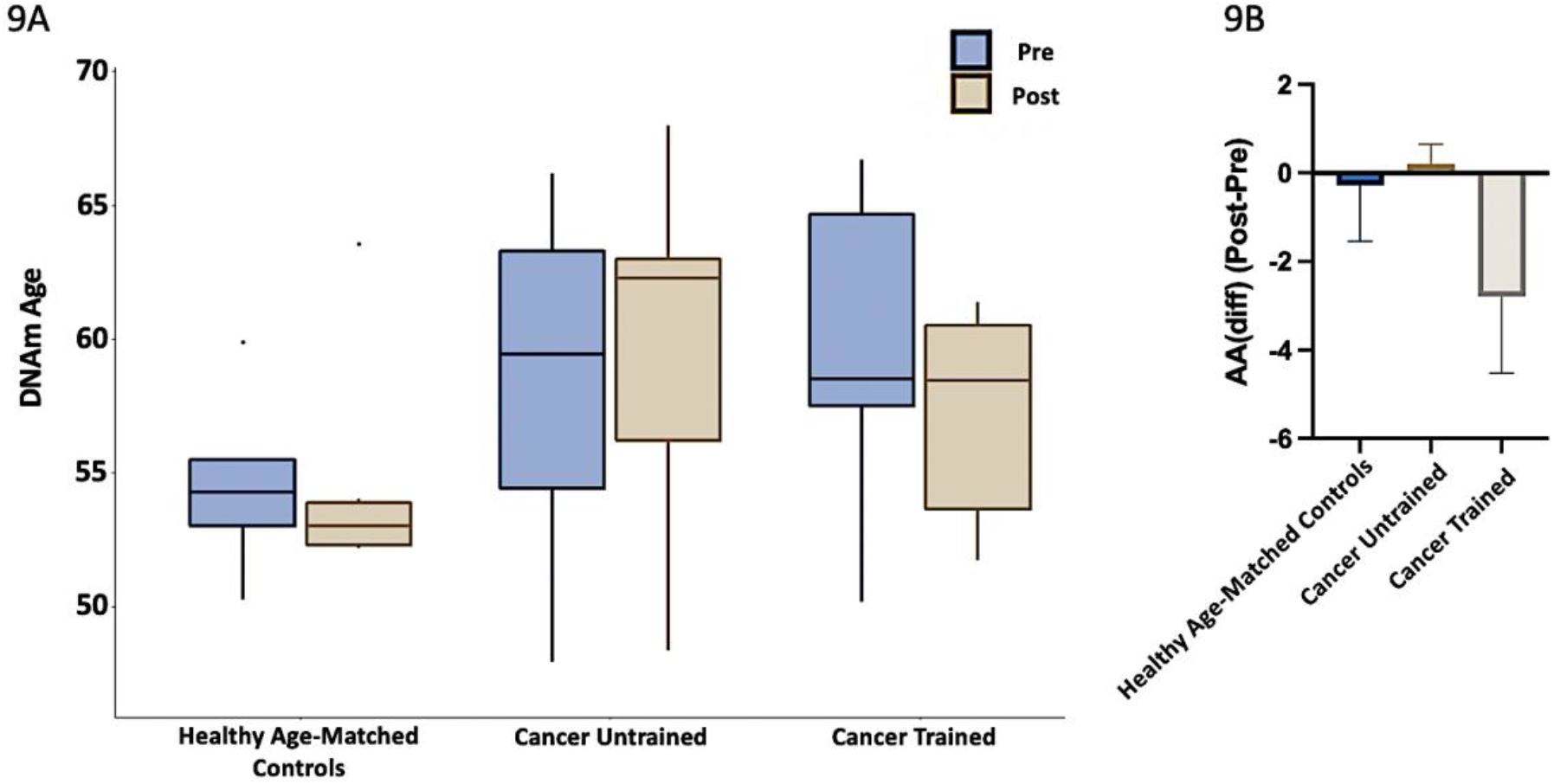
**A**. The muscle specific epigenetic age test (MEAT) that estimates DNA methylation (DNAm) Age using the MEAT2.0 elastic net regression model [31, 32]. From left to right, plots show DNAm Age in the: 1) Healthy age-matched trained control group pre and post aerobic training; 2) in the cancer untrained group pre and post aerobic training, and; 3) in the cancer trained group pre and post aerobic training. Boxplots are median and inter quartile range (IQR). The lower whisker extends from the 25 percentiles to the lowest value within 1.5* IQR, while the upper whisker extends from the 75 percentiles to the highest value within 1.5* IQR. Outliers (as specified by Tukey) are plotted as points beyond the end of the whisker. **B** Age acceleration (AA) calculated by DNAm-Age minus chronological age and the difference between post vs. pre denoted by AA(diff) (Post-Pre).

## Discussion

In line with our original hypotheses, we were able to demonstrate that there was retained DNA methylation in the SkM of cancer survivors even 10-14 years after diagnosis and treatment compared with healthy (non-cancer) age-matched controls. Aerobic training helped ‘rejuvenate’ or reset some of the epigenetic landscape evoked by cancer treatment and survival back towards DNA methylation signatures observed in healthy age-matched controls. Also, on average, cancer survival evoked a higher SkM epigenetic age compared to healthy age-matched controls, however this was not statistically significant. Further, there was no reduction in epigenetic age in cancer survivors following training. On average, the training could also prevent a further increase in epigenetic age versus cancer survivors who did not undertake any aerobic training, however this was not statistically significant.

### Retained DNA methylation in the skeletal muscle of cancer survivors even 10-14 years after diagnosis and treatment

We and others have previously demonstrated that aged individuals possess hypermethylated DNA in their SkM [27-29, 31, 32]. We have also demonstrated that human muscle tissue retains prolonged epigenetic (DNA methylation) signatures following a period of exercise induced hypertrophy [17, 18] and more recently this has been confirmed in mice [19]. However, these epigenetic memory imprints have only been measured after total of 22 weeks following the first bout of exercise in humans. Here we demonstrate that cancer survivors, even 10-14 years after diagnosis and treatment from breast cancer demonstrate increased hypermethylation in SkM specifically in gene regulatory regions of CpG islands within promoters compared to healthy age-matched controls. Where 98% of the significantly differentially methylated DNA sites in CpG islands within promoter regions were hypermethylated compared with only 2% that were hypomethylated. Hypermethylation in these gene regulatory regions was enriched in: Ubiquitin Mediated Proteolysis (UPS), Cell Cycle, Mitophagy, Autophagy and p53 pathways. Hypermethylation of these pathways would suggest a suppression of genes within these pathways. Indeed, a reduction in gene expression associated with cell cycle could impair growth and normal muscle maintenance (reviewed in [50]). Therefore, hypermethylation and suppression of gene expression within proteolysis, mitophagy and autophagy pathways, while perhaps counterintuitive as increases in these processes are observed in muscle wasting conditions such as cancer cachexia [51-53], this data is perhaps suggestive of a dysregulation in the genes involved in protein quality control processes in the SkM of cancer survivors. Where an inability to properly remove damaged cellular and mitochondrial proteins could contribute to impaired muscle function and quality [53, 54]. For example, blocking autophagy is unable to spare muscle mass in mice with cancer cachexia, due to its role in protein quality control [55]. And studies have recently suggested that it is the continued ability of the muscle to undergo autophagy and mitophagy in an efficient manner that can help maintain mass and myofiber integrity, where inhibition of these important processes can contribute to the accumulation of abnormal mitochondria, muscle degeneration and weakness [54].

One other potential contributing factor to the retained epigenetic signatures in cancer survivors is that these individuals may have had different lifestyle choices post diagnosis and treatment, such as the amount of physical activity, compared to healthy individuals. However, we confirmed that the reported physical activity levels between cancer survivors and healthy age-matched control group were not significantly different, and probably not a likely cause for the retained hypermethylated profiles observed. Even if this was the case, increased physical activity has been shown to promote hypomethylation [28, 39] not hypermethylation that was observed in the cancer survivors. The other lifestyle choice that could have been altered between the cancer survivors and the healthy age-matched controls is their diet. Where for example, a high fat diet can promote hypermethylation of SkM [21]. While we did not gather information on habitual dietary intake between these groups, it is perhaps more logical due to being diagnosed and treated for cancer, this groups diet may have been healthier than the age-matched controls, and that would perhaps therefore have a positive impact on the methylome. Where positive encounters (such as exercise-discussed below) in muscle tend to be a hypomethylating, not hypermethylating stimulus that was observed in cancer survivors after aerobic training. However, at present the role of long-term alterations in diet and nutrient intake in SkM and the retention of epigenetic imprints across the genome are lacking and requires further investigation.

### Aerobic exercise training in cancer survivors helped ‘rejuvenate’ or reset some of the epigenetic landscape towards DNA methylation signatures observed in the healthy

We and others have demonstrated that aerobic, high-intensity and resistance exercise is a hypomethylating (and predominantly gene turning on stimuli) in SkM [17, 18, 33-35]. With both acute exercise and chronic training identified to promote hypomethylation in human and mouse SkM [17, 18, 30, 33-35]. Indeed, acute aerobic, sprint interval [34] and resistance exercise [17, 18], as well as chronic resistance training in humans [17, 18] and chronic weighted wheel running training in mice [19, 29] all demonstrate these enriched hypomethylated signatures. Albeit, the different exercise types (e.g. aerobic [35] vs. resistance [17, 18] vs. high intensity exercise [34]) promote this hypomethylation of divergent genes and pathways. Exercise has therefore been proposed to be epigenetically ‘anti-ageing’ [28, 29, 36-39]. Where we have also demonstrated that resistance exercise can reset the mitochondrial methylome in elderly individuals [36] and others have demonstrated this in aged humans and mice where resistance exercise and power weighted wheel running, respectively, can help return DNA methylation signatures back towards those observed in younger individuals [29, 37]. Indeed, total higher physical activity levels in the elderly is also associated with younger epigenetic profiles [39] and younger individuals with higher activity levels of aerobic exercise demonstrate hypomethylation of the same genes that are hypermethylated in the SkM of elderly people [28]. However, no one has investigated the methylome of cancer patients following a period of aerobic training.

Therefore, the present study was the first to demonstrate the hypomethylating effect of aerobic training in gene regulatory regions in cancer survivors. Where, aerobic training elicited a considerable hypomethylation in 99% of the DMPs located specifically in CpG islands within promoter regions. Importantly, training was able to reverse the hypermethylation identified in cancer survivors back towards a hypomethylated signature that was observed pre-training in healthy age-matched controls at 300 (out of 881) of these island and promoter associated CpG sites. Pathway enrichment analysis identified that the training in cancer survivors evoked this predominantly hypomethylated signature in: Cell cycle, DNA replication and repair, transcription, translation, mTOR signalling and the proteosome pathways. Interestingly, as discussed above, cell cycle was also demonstrated to be hypermethylated in the cancer survivors compared to healthy individuals at baseline. Therefore, the aerobic training in cancer survivors was able to reverse the profile in many cell cycle genes towards a hypomethylated profile. Indeed, as discussed above, cell cycle genes have fundamental processes in SkM, particularly for muscle stem cell (satellite cell) activation, fusion/differentiation, and quiescence to maintain the muscle stem cell pool with age (reviewed in [50]). Therefore, a move from hypermethylation in cancer survivors at baseline toward a hypomethylated signature after training would potentially increase cell cycle-related gene expression, leading to improved growth-related processes in SkM of cancer survivors. It still remains to be determined how long these hypomethylated signatures could be retained after aerobic training is ceased.

Differentially methylated region (DMR) analysis also identified 11 genes in CpG islands within promoter regions as hypermethylated in breast cancer survivors with training reversing these promoter associated DMRs towards a hypomethylated signature. Most of these genes were associated with the proteosome (BAG1, SPG7), cell cycle / mitosis / DNA replication and repair (BTG2, CHP1, KIFC1, POLD2, S100A6), RNA transcription (MKL2, SNORD104) and peroxisomal proliferation (PEX11B). We confirmed that these genes had multiple sites in their promoter regions that were hypermethylated in breast cancer survivors and then demonstrated a reversal to hypomethylation after aerobic training. This is significant as these genes are also primarily a part of the enriched pathways that, as previously mentioned, demonstrated hypermethylation in cancer survivors. A particularly interesting DMR we identified within the promoter region of the MTR-gene, as MTR is responsible for the regeneration of methionine from homocysteine. Where the generation of methionine is critical for the synthesis of S-adenosylmethionine (SAM), with SAM then acting as a co-factor for DNMT (DNA methyltransferase) which is the universal methyl group donor for the process of DNA methylation itself. It is therefore interesting that a region within this gene’s promoter region is hypermethylated in breast cancer survivors and hypomethylated after aerobic training. We were also able to confirm that hypermethylation in breast cancer survivors led to significant reductions in MTR gene expression. The hypomethylation with training in cancer survivors was also able to increase average MTR gene expression across the group, however, this was not statistically significant as some individuals increased MTR expression with training and some also reduced expression. We were also able to demonstrate that the hypermethylation of the DMRs after cancer survival, demonstrated significantly lower gene expression in cancer survivors compared with healthy individuals at baseline for the majority of genes. Furthermore, several of these DMRs then demonstrated a reversal to hypomethylation after training in cancer survivors and also increased gene expression. However, none of these genes demonstrated statistically significant increases with training. This may be due to the varied response across cancer survivors, with some individuals increasing gene expression, some remaining unchanged and a small number of individuals demonstrating reduced gene expression. Therefore, the alterations in gene expression following training in cancer survivors demonstrated quite an individualised response. This may reflect, that while exclusion criteria aimed to recruit breast cancer survivors with similar backgrounds, these individuals would still have received varied treatment regimens that may affect their ability to respond to the training stimulus that is perhaps not due to epigenetic or transcriptional changes, rather the activity of signal transduction networks at the protein level. However, it was clear that the cancer survival significantly hypermethylated and reduced the expression of these genes compared with healthy controls, indicating that perhaps the abundance of these genes at the protein level would have been reduced to begin with prior to commencing aerobic training.

A gene that is worth highlighting in this study is BTG2 (BTG Anti-Proliferation Factor 2). There was a DMR located in BTG2’s promoter region that was hypermethylated after cancer survival and reverted to a hypomethylated profile after training in cancer survivors. This gene also demonstrated significant higher gene expression in cancer survivors at baseline compared with healthy individuals, switching to a significant decrease in gene expression after training in cancer survivors. At the gene and protein level this would perhaps make sense given BTG2’s proposed role as an antiproliferative protein, where it can inhibit the CCR4-NOT complex involved in cell cycle regulation [56]. Further, BTG2 can impair cardiac hypertrophy via binding to proteins that comprise mRNA deadenylation complexes, therefore suppressing cytosolic RNA levels [56]. Therefore, increases in BTG2 gene expression in the muscle after cancer survival could be hypothesised to impair growth and transcription and decreases expression after training could perhaps improve growth and transcription levels in muscle tissue.

Finally, it is also worth pointing out that there was very little overlap between the methylation changes seen in both DMPs, DMRs and across pathways after aerobic training in cancer survivors and after the same exercise training program in healthy individuals. This further confirms that cancer survivors had very different DNA methylation profiles in their SkM prior to training, and that the epigenetic remodelling of the genome following training was also vastly different. The only gene to demonstrate DMPs that were altered with exercise regardless of disease/health background was SIRT2. SIRT2 is a histone deacetylase where its inhibition reduces muscle mass and endurance performance [57, 58], and its deletion impairs regeneration after SkM injury [57]. SIRT2 also regulates cell cycle, autophagy and DNA repair (e.g., via FOXO). With these pathways identified above as hypermethylated with cancer survival and hypomethylated after aerobic training. Interestingly, SIRT2 was differentially regulated, where aerobic training in cancer survivors elicited a hypomethylating effect compared to a hypermethylation effect in healthy individuals. Therefore, the data from the present study suggests that SIRT2 gene is differentially methylated after both disease (cancer) and aerobic training regardless of disease history (i.e., in both cancer and healthy individuals) but in an alternate fashion in cancer survivors compared with healthy individuals. We also confirmed that gene expression of SIRT2 was reduced in cancer survivors compared to healthy age-matched controls. However, aerobic training in both groups, on average, increased SIRT2 (albeit non-significantly) therefore the divergent epigenetic signature of SIRT2 in response to training in the healthy vs. cancer survivors was not translated to alterations at the gene expression level.

Overall aerobic training helped ‘rejuvenate’ or reset some of the epigenetic landscape evoked by cancer survival back towards DNA methylation signatures observed in healthy age-matched controls. This occurred particularly in gene regions associated with cell cycle, mitosis, transcription and the proteosome (including autophagy and mitophagy) and aerobic training evoke a considerably different epigenetic signature in cancer survivors compared with healthy individuals.

### Cancer survival evokes an average increase in epigenetic age in skeletal muscle

The epigenetic modification of DNA methylation has been shown to form an epigenetic clock that can predict both chronological and biological age [22-24]. Indeed, epigenetic age using DNA methylation from whole blood has been shown to increase in breast cancer patients undergoing treatment [25]. A recent study in childhood cancer, suggested that these individuals demonstrated accelerated epigenetic age into adulthood [26]. However, the role of exercise/aerobic training’s ability to alter increased epigenetic ageing after cancer survival is unknown. We have recently developed a muscle-specific epigenetic clock (muscle-specific epigenetic age test / MEAT) that predicts age with better accuracy than the pan-tissue epigenetic clock [31, 32]. Using this epigenetic clock, we identified that on average cancer survival resulted in higher epigenetic age in SkM compared to healthy age-matched controls. Further, while there was no reduction in epigenetic age in cancer survivors following aerobic training, on average the training could prevent a further increase in epigenetic age versus cancer survivors who did not undertake any aerobic training, however, did not reach statistical significance. Further when calculating DNA methylation age acceleration, after training in cancer survivors there was a trend for a reduction in age acceleration that was not altered in the healthy age-matched controls or untrained cancer survivors. Suggesting a trend towards decelerated DNA methylation age in cancer survivors following training. However, this difference was also not statistically significant between groups. The lack of statistical differences in the epigenetic age data was probably due to interindividual variability in this data, where some cancer survivors reduced their epigenetic age due to training whereas others showed no change. Therefore, a larger sample size was probably required to perform this epigenetic age analysis in this study.

## Conclusion

Even 10-14 years after breast cancer diagnosis, treatment, and survival, human SkM possessed retention of hypermethylated DNA signatures compared with healthy age-matched controls. Importantly, aerobic training helped rejuvenate the human SkM methylome towards signatures identified in healthy age-matched individuals in gene regulatory regions.

## Data Availability

Raw and normalised DNA methylome array data will be deposited and freely available on deposited on GEO https://www.ncbi.nlm.nih.gov/geo/ under accession number: GSE213029. All analysed data is included as supplementary files in this manuscript.

## Acknowledgements and funding statement

The study is funded by the Norwegian Cancer Society and Aktiv mot kreft (Active Against Cancer). The study has used data from the Cancer Registry of Norway. The interpretation and reporting of these data are the sole responsibility of the authors and no endorsement by the Cancer Registry of Norway is intended nor should be inferred. Adam P. Sharples’ group is funded by the Research Council of Norway (grant 314157).

## Data Availability Statement

DNA methylome array data will be deposited and freely available on deposited on GEO https://www.ncbi.nlm.nih.gov/geo/ under accession number: GSE213029.

