## Supplementary material for "Aerobic Exercise Training Rejuvenates the Human Skeletal Muscle Methylome Ten Years after Breast Cancer Treatment and Survival": Suppl. Figure 1

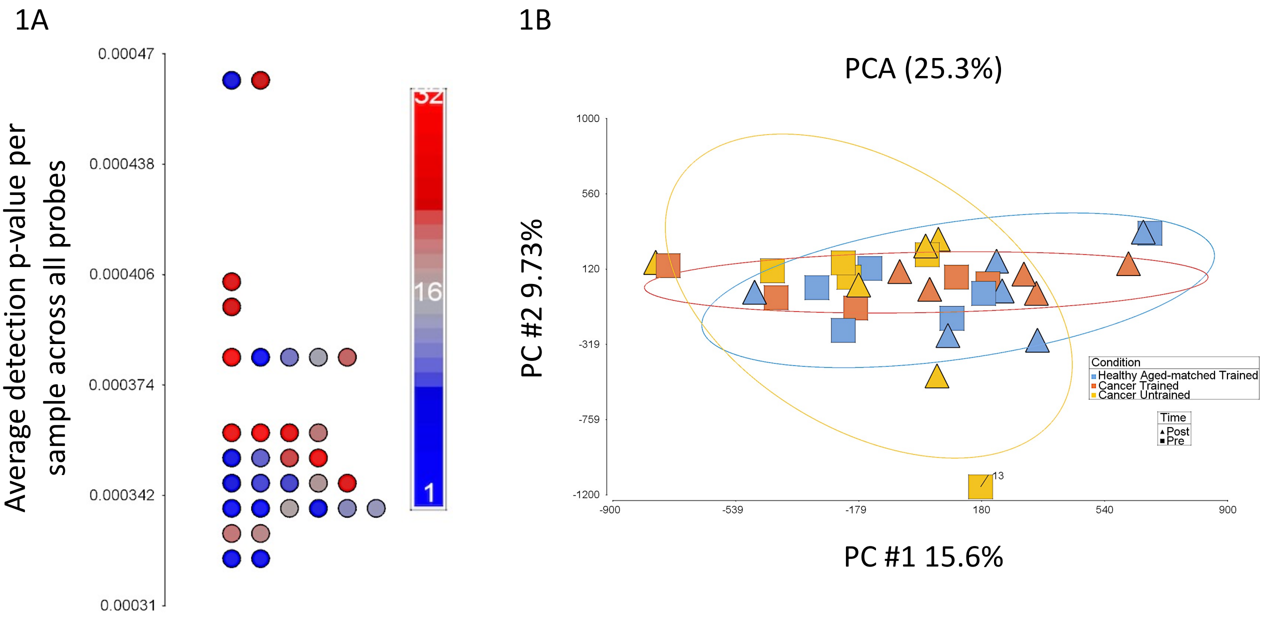


**Suppl. Figure 1. A.** Average detection p-value per sample across all probes, with all samples with values less than 0.0004. **B.** PCA for all individual samples across all conditions coloured by condition/group (Cancer Trained- orange, Cancer Untrained- yellow and Healthy Aged-matched Controls- blue) and shaped by time (pre and post training, squares and triangles respectively). One sample (sample 13) was removed due to a larger variation than that expected within that condition/group (defined as values above 2.2 standard deviations for that condition).
